# Epigenetic age deviation and mental health in childhood and adolescence: A systematic review and meta-analysis

**DOI:** 10.1101/2024.05.06.24306916

**Authors:** L.B. Moyakhe, S. Dalvie, T.C. Chalumbila, D.J. Stein, N. Koen

**Affiliations:** Department of Psychiatry and Mental Health, Faculty of Health Sciences, University of Cape Town, South Africa; and UCT Neuroscience Institute; South African Medical Research Council (SAMRC) Unit on Risk and Resilience in Mental Disorders; SAMRC Genomic and Precision Medicine Unit, Division of Human Genetics, Department of Pathology, Institute of Infectious Diseases and Molecular Medicine, University of Cape Town□; Department of Biomedical Sciences, Faculty of Medicine, University of Botswana

## Abstract

**Background:** Epigenetic age acceleration (EAA, i.e., higher EA relative to chronological age) may be linked to adverse mental health outcomes in children. Previously, EAA has been associated with advanced physical maturation and early pubertal development in adolescents. However, research on epigenomic changes and mental health outcomes in children remains limited. This systematic review aimed to investigate the associations between epigenetic age deviation (greater or lower EA age relative to chronological age) and mental health outcomes in childhood.

**Methods:** This systematic review adhered to the Preferred Reporting Items for Systematic Reviews and Meta-analysis (PRISMA) guidelines. Relevant terms were used to search the PubMed, Scopus, and PsychINFO (via Ebsco host) online databases. The search commenced between February and September 2022. Only full-text studies published in English, involving participants under 18 years of age, and examining associations between epigenetic age deviation and child mental health outcomes were eligible for inclusion.

**Results and conclusion:** Among the 4 studies that met the inclusion criteria for this review, 3 studies independently reported significant associations between EAA and internalising behaviour in children and adolescents (aged 4 to 17 years). However, a meta-analysis (OR=1.14, [95% CI, 0.86-1.49]) incorporating a subset of these studies (n=2) did not confirm this finding, though heterogeneity between studies was observed (I^2^ =81%, p=0.022). While the data are not consistent, the findings of this systematic review suggest that EAA holds promise as a potential biomarker for identifying children and adolescents at risk of internalising problems. Given the limited number of studies and the heterogeneity in effect size measures, further work is warranted to explore these preliminary findings.

## Introduction

Mental health disorders – including internalising behaviours (such as anxiety and depression) and externalising behaviours (such as disruptive behaviours and attention deficit hyperactivity disorder (ADHD)) - often emerge during childhood and adolescence (Kessler et al., 2007, Shaw et al., 2012). These disorders may lead to disrupted developmental trajectories and may precede adverse educational, social, and occupational outcomes that persist into adulthood (Shaw et al., 2012, Kieling et al., 2011, Sallis et al., 2019). Prior studies have also shown that risk for mental disorders during childhood and adolescence may be associated with advanced physical ageing (e.g. early pubertal timing) (Copeland et al., 2019).

Epigenetic mechanisms, i.e. DNA methylation (DNAm) have been implicated as potential risk factors for mental health outcomes. For example, Barker and colleagues showed that a methylation index for inflammation risk correlated with internalising behaviours during childhood (Barker et al., 2018). Moreover, indicators of advanced biological ageing based on genomic DNAm have become of interest and have emerged as a cross-tissue index of biological ageing (Horvath and Raj, 2018). Epigenetic clocks utilise DNAm values at selected CpG sites (determined through machine learning approaches) to provide estimates of age across various tissues at different stages of life (Horvath, 2013, Jung et al., 2017). These estimates, referred to as epigenetic age (EA, or DNAmAge), have been found to predict age-related phenotypes (Horvath and Raj, 2018). Increased epigenetic age relative to chronological age (i.e., EA acceleration (EAA)), has been associated with adverse health outcomes, including cardiovascular disease and mortality in adults (Fransquet et al., 2019).

Emerging evidence also suggests potential associations between EAA and mental health disorders in adults, such as depression and bipolar disorder (Han et al., 2018, Fries et al., 2017). Similarly, EAA was found to be associated with early stress exposure in children, which in turn was positively linked to depressive behaviours in adolescence (Sumner et al.,2019). EA deceleration has also been associated with lower developmental maturity in children (Girchenko et al., 2017, Knight et al., 2018). However, studies evaluating early biomarkers that may link environmental factors to mental health outcomes in children are currently limited (Cerveira de Baumont et al., 2021). Moreover, there has been relatively minimal work investigating EA deviation (i.e. higher or lower EA) during childhood (Dieckmann et al., 2021). Therefore, we aimed to systematically review the existing literature on the associations between EA deviation and adverse developmental and mental health outcomes in childhood and adolescence. To the best of our knowledge, this was the first such review undertaken.

## Methods

This review was conducted per the Preferred Reporting Items for Systematic Reviews and Meta-Analyses (PRISMA) guidelines for protocol, search strategy, and risk of bias assessment (Moher et al., 2009); and subsequently registered on the International Prospective Register of Systematic Reviews (PROSPERO CRD42022348873). A comprehensive electronic database search was conducted in PubMed, Scopus and PsychINFO via Ebscohost, between February and September 2022. The search terms included “neurodevelopment disorders,” “developmental psychopathology,” “internalizing disorder,” “externalizing disorder,” “behaviour disorder,” “youth,” “epigenetic age” and “biological ageing”. Full details of the search strategy and search terms are provided in Appendix 1, **Supplementary Material**.

### Study selection criteria

Following the comprehensive search, title and abstract screening were conducted according to the inclusion criteria. Full-text studies published in English and evaluating epigenetic age deviation were eligible for inclusion. Additionally, only studies of participants younger than 18 years old were eligible for inclusion (with no further demographic restrictions). Epigenome-wide association studies (EWAS) without an EA deviation component and/or not assessing developmental or mental health outcomes in childhood/adolescence were excluded.

### Quality assessment

Two independent reviewers (L.B Moyakhe and T.C Chalumbila) conducted a quality assessment of the studies using Q-Genie, a quality assessment tool designed for genetic studies (Sohani et al., 2016). Potential sources of bias and appropriateness of sample sizes were also assessed. Each item was marked on a 7-point Likert scale – studies were rated as moderate-quality if scoring a total of >32 but ≤40 on Q-Genie; and good-quality if scoring > 40. Complete details of the quality assessment ratings for each included study are provided in Appendix 2, **Supplementary Material**.

### Data extraction

Data were extracted independently by the two reviewers (L.B Moyakhe and T.C Chalumbila) using a data extraction form developed for this review, Appendix 2, **Supplementary Material**. Information extracted included study design, location, and sample characteristics; biological sampling; the measure (epigenetic clock) used to determine EA; the outcome phenotype (e.g., internalising disorders); the outcome measure (e.g., Child Behaviour Checklist (CBCL)); and the main study findings. The elimination process involved the removal of duplicates and the appraisal of study titles and abstracts to exclude non-relevant studies.

### Meta-analysis

A random-effects meta-analysis was performed using the *metafor* package in R (Viechtbauer, 2010). Specifying random-effects accounts for heterogeneity in the true associations which are attributable to factors that may lead to sample variation across cohorts (e.g. differences in measurements or sample characteristics). This meta-analysis was undertaken using odds ratios (OR) and confidence intervals (CI). A forest plot was also generated.

## Results

### Search results

Study selection was undertaken by the first author (L.B. Moyakhe). A total of 1200 studies were retrieved; 52 were duplicates and subsequently removed, **Figure 1**. Title and abstract screening were then performed on 1148 studies, after which only 8 studies were found to be eligible for full-text screening. Full-text screening and quality assessment then yielded 4 eligible studies, from which data were extracted and included for further analysis. Excluded studies were those with adult participants; those with non-DNAm EA measures (e.g., telomere length); and those not assessing the outcomes of interest.

**Figure 1.**
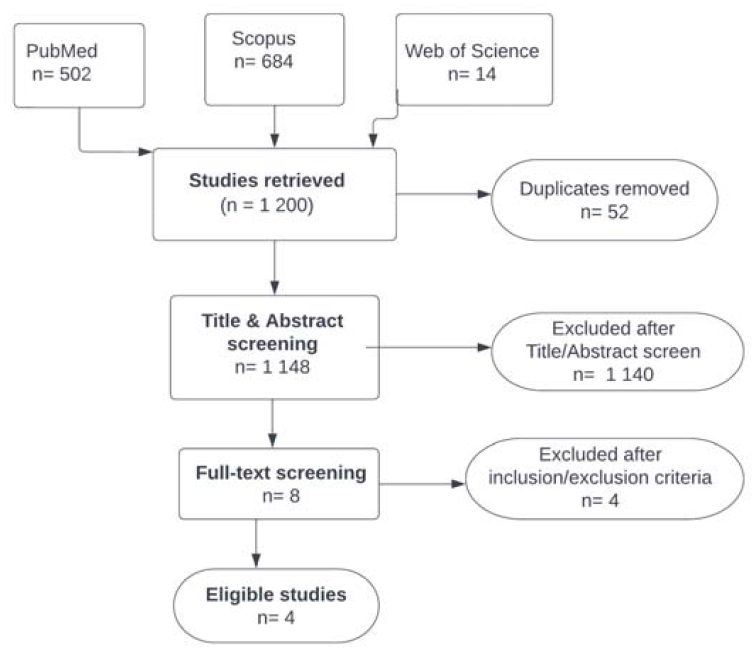
PRISMA flow diagram of study selection.

### Study and sample characteristics

A detailed description of the included studies (n=4) is presented in **Table 1**. Per the inclusion criteria for this review, all study participants were ≤18 years of age. Three of the studies utilised the Horvath epigenetic clock (Suarez et al., 2018a, Tollenaar et al., 2021, Cerveira de Baumont et al., 2021), a widely recognised measure of epigenetic ageing (Horvath, 2013); while the fourth study employed the Pediatric Buccal Epigenetic clock (PedBE) (Cerveira de Baumont et al., 2021), specifically designed for use with children (McEwen et al., 2020).

**Table 1.** Characteristics of included studies – epigenetic ageing and developmental and mental health outcomes in children.

| Reference (PMID) | Target cohort | Age (years) | Sample Size | Ancestry | Relevant Findings | Outcome Phenotype | Outcome Measure | Epigenetic clock | Biological Sample |
| --- | --- | --- | --- | --- | --- | --- | --- | --- | --- |
| Suarez et al., 2018<br>(30021623) | Glycyrrhizin in Licorice (GLAKU) | 12.3 | N=239 | Caucasian | - EAA assoc with 18 to 34% higher odds for internalizing and thought problems<br>- EAA was not significantly associated with cognition. | Internalising problems<br>Externalising problems | Maternal report CBCL (6-18)<br>The short form of the WISC-III for cognitive function | Horvath | Blood |
| Tollenaar et al., 2021<br>(33460784) | Longitudinal Basale Invloeden Op De Baby Ontwikkeling (BIBO) study | 6 - 10 (longitudinal) | N=148 | Caucasian | - EAA assoc with internalising symptoms at 6 to 10<br>- Internalising symptoms at 2.5 and 4 years predicted EAA at age 6. | Internalising symptoms<br>Externalising symptoms<br>EAA at age 6 | Maternal report CBCL 1.5-5 (at ages 2.5 and 4)<br>CBCL 4-28 (at ages 6, 7 and 10)<br>Follow-up: SDQ (ages 8 10) | Horvath | Buccal epithelial cells |
| CerveiradeBaumont et al., 2021<br>(33833304) | Children and adolescents were recruited from public schools in Brazil | Baseline 13.4<br>Follow-up 17.2 | N=234, (N=76 after 5-year follow up) | Caucasian 63%,<br>*African Brazilian 14%,<br>*Mixed 21% | - No difference in EA between adolescents with/without an anxiety disorder. | Anxiety | Initial diagnoses: psychiatric evaluation<br>Follow-up: K-SADS-PL <18 | Horvath | Saliva |
| Dammering et al., 2021<br>(34621920) | Berlin Longitudinal Children Study | 4.25 | N=158 | White: 144 (91 %)<br>*White-black: 11 (7%)<br>*White-Asian: 3 (2 %) | - Children with internalizing disorder exhibited significant EAA compared to children without internalizing disorder. | Internalising disorders | DSM-IV criteria using the electronic Preschool Age Psychiatric Assessment. | Pediatric buccal epigenetic (PedBE) clock | Saliva |

- CBCL=Child Behaviour Checklist (versions, versions 1.5-5; 4-28; 6-18); K-SADS= Schedule for Affective Disorders and Schizophrenia for School-Age Children Present and Lifetime WISC-III= Wechsler-Intelligence-Scale-for-Children-III; SDQ= Strengths and Difficulties Questionnaire; DSM-IV= Diagnostic and Statistical Manual of Mental Disorders fourth edition.
\*Ancestry described as in the original study
\*Ancestry % not reported by study but reported for consistency
Note: With the exception of Tollenaar et al (2021), ages are reported from studies as mean ages

The prospective cohorts included in each of these four studies were diverse, spanning a number of geographical locations and community settings: The Glycyrrhizin in Licorice (GLAKU), a community-based cohort in Finland (Strandberg et al., 2002); the Longitudinal Basale Invloeden Op De Baby Ontwikkeling (BIBO) study in the Netherlands (Beijers et al., 2010); children recruited from public schools in Brazil (Salum et al., 2011); and the Berlin Longitudinal Children Study (Heim, 2017). The total sample size across all four studies was approximately 770 participants. The majority of these studies included participants of European ancestry (Tollenaar et al., 2021; Dammering et al., 202; Suarez et al., 2018), except for one study which included participants of diverse ancestry (e.g. mixed and African Brazilian participants) (Cerveira de Baumont et al., 2021). In terms of outcome measures, three of the included studies assessed internalising behaviour (Tollenaar et al., 2021; Dammering et al., 2021b; Suarez et al., 2018b), and of these, two studies also assessed externalising behaviour (Tollenaar et al., 2021; Suarez et al., 2018a). The fourth study assessed anxiety disorders (Cerveira de Baumont et al., 2021).

### Quality assessment

The quality assessment of the included studies was conducted using the Q-Genie checklist (Sohani et al., 2016). All the studies included in this review were rated as good quality (i.e., with scores >40). The main limitations identified based on the Q-genie checklist were limited sample size and reduced statistical power across the studies. Due to heterogeneity in the effect size measures reported by the individual studies, conducting a meta-analysis was only feasible for two of the four included studies.

### Associations between epigenetic age (EA) deviation and internalising behaviour

Associations between EA deviation and internalising behaviour were examined in three studies involving children aged 4 to 13 years (Suarez et al., 2018a, Tollenaar et al., 2021, Dammering et al., 2021). Internalising behaviour was assessed using the CBCL (versions 1.5-5; 4-28; 6-18) (Suarez et al., 2018a; Tollenaar et al., 2021), and the electronic Preschool Age Psychiatric Assessment (Bell, 1994) (Dammering et al., 2021), based on the Diagnostic and Statistical Manual of Mental Disorders, fourth edition (DSM-IV) (Guze et al.,1995). The individual study sample sizes were relatively small, ranging from 150 to 240 participants per study. While most participants were of European ancestry, two studies also included participants of diverse ancestry – _such as mixed ancestry and African Brazilian (Cerveira de Baumont et al., 2021, Dammering et al., 2021). Overall, all three studies reported associations between EAA and internalising behaviour (Suarez et al., 2018a; Tollenaar et al., 2021; Dammering et al., 2021a). Notably, only one study utilised the PedBE clock (Dammering et al., 2021), and the remaining two studies utilised the Horvath clock as the measure of EA (Suarez et al., 2018a; Tollenaar et al., 2021). Thus, these two studies utilise the same epigenetic clock and the same effect size estimates - reporting that EAA was associated with increased odds for internalising behaviour in children (OR=1.01, CI (0.98-1.08)) (Tollenaar et al., 2021); (OR= 1.34, CI (1.10-1.58)) (Suarez et al., 2018a) - were therefore eligible for meta-analysis.

### Associations between EA deviation and anxiety disorders

One study examined the association between EAA and anxiety disorder (Cerveira de Baumont et al., 2021). In this study, the baseline assessment was conducted at about 13 years of age, with a follow-up assessment at 17 years old. Initial diagnoses were obtained from psychiatric evaluations and the Schedule for Affective Disorders and Schizophrenia for School-Age Children Present and Lifetime Version (K-SADS-PL) (Kaufman et al., 1997) for participants under 18 years old. This study’s findings reported no significant difference in EAA between adolescents with and without anxiety disorders (Cerveira de Baumont et al., 2021).

### Associations between EA deviation and externalising behaviour

Two studies examined associations between EA and externalising behaviour in children aged 2 to 13 years (Suarez et al., 2018a; Tollenaar et al., 2021). Both studies used the CBCL as the assessment measure; and neither study reported any significant associations between EA and externalising behaviour (Suarez et al., 2018a; Tollenaar et al., 2021).

### Meta-analysis of EA and internalising behaviour

A random-effects meta-analysis was conducted on two of the four included studies, which investigated the associations between EAA and internalising behaviour (Suarez et al., 2018; Tollenaar et al., 2021). In contrast to the findings of the individual studies, the meta-analysis yielded no significant difference between EAA and internalising behaviour (OR=1.14, [95% CI, 0.86-1.49]), **Figure 2**. However, there was evidence of significant heterogeneity between the studies (I^2^ = 81 %, p-value = 0.022). A meta-analysis of the remaining two studies was not performed due to heterogeneity in the effect-size measures.

**Figure 1.**
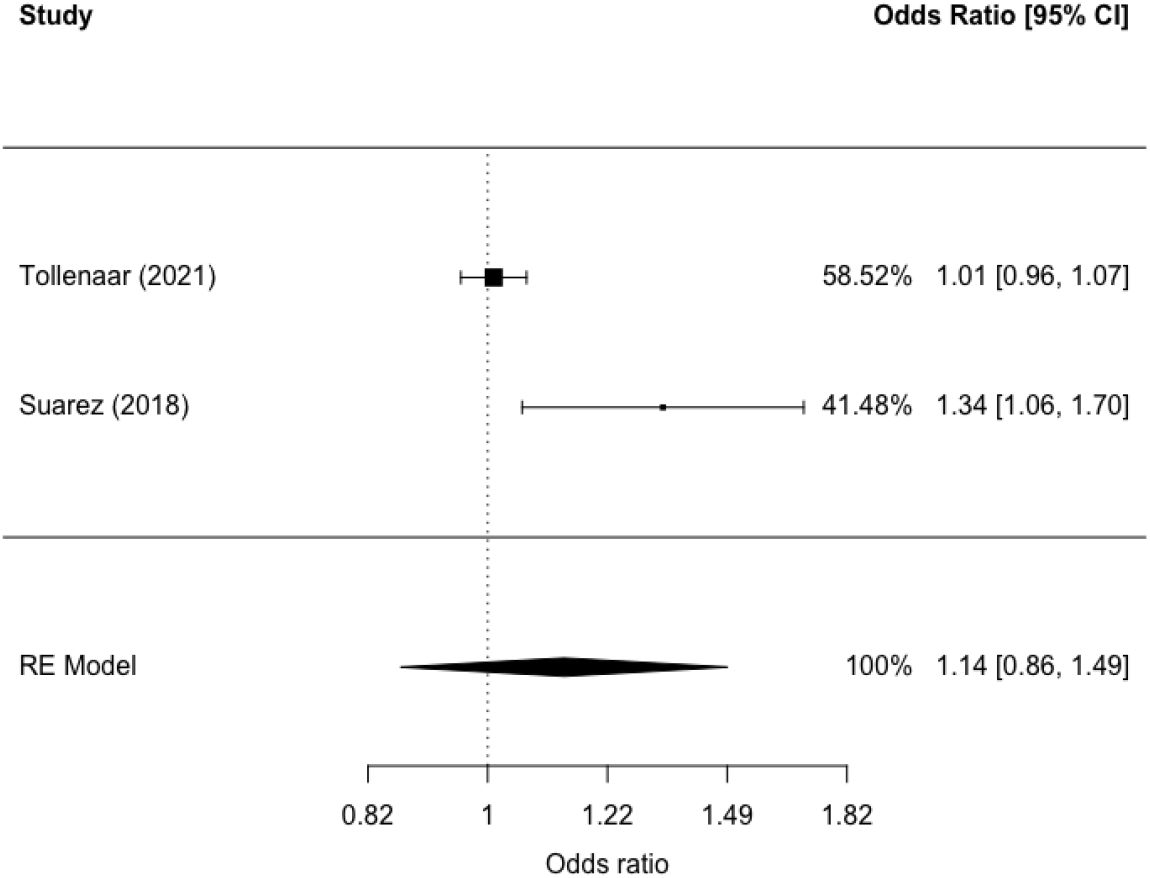
Random-effects meta-analysis forest plot of the associations between epigenetic age acceleration (EAA) and internalising behaviour in children. The bars represent confidence intervals corresponding to α=.05. The % represents the weights of the study. Model p-value= 0.36, OR=1.14, CI (0.86 -1.46). Test for heterogeneity I^2^= 81 %, p-value = 0.022.

## Discussion

This systematic review collated findings from four studies investigating the associations between EA deviation and mental health outcomes in childhood and adolescence. EAA was significantly associated with internalising behaviour in children in three of the four studies (Suarez et al., 2018a, Tollenaar et al., 2021, Dammering et al., 2021). For example, one study found EAA associated with an increased risk for internalising behaviour (Suarez et al., 2018a). Similarly, children with an internalising disorder diagnosis had higher EAA when compared to those without such a diagnosis (Dammering et al., 2021). In this study, children at risk for internalising behaviour are also associated with EAA and physical growth (Dammering et al., 2021). These findings are consistent with previous literature linking EA with physical maturation in children (Copeland et al., 2019; Ellis et al., 2019). Further, prior studies reported associations between EAA and adverse outcomes such as internalising symptoms (e.g. depressive symptoms) in childhood and late adolescence (ages 15 to 17) (Suarez et al., 2018b, Caro et al., 2023).

However, the meta-analysis conducted on a subset of the studies (n=2) in this systematic review did not confirm the findings observed in the individual studies. Substantial heterogeneity (e.g. in study settings or individual participant characteristics) was observed in the random-effects meta-analysis. Thus, considerable variation across the included studies may have contributed to the non-significant findings. This would align with prior evidence that the association between EAA and the development of internalising behaviours is both dynamic and heterogeneous within any given population (Caro et al., 2023, Barker and Maughan, 2009). For example, one study reported that EAA at birth is associated with lower internalising behaviours in early childhood for some children, while potentially higher internalising behaviours at the start of adolescence for others (Caro et al., 2023). Notably, one study in this review also reported no significant associations (i.e. between EAA and anxiety disorders in adolescence) (Cerveira de Baumont et al., 2021), thus aligning with the findings of the meta-analysis. This is consistent with previous literature in adults reporting no observable differences in EAA between patients with generalised anxiety disorder and healthy controls (Wolf et al., 2019).

This review has several noteworthy limitations. First, the search strategy and study selection process may have resulted in the omission of relevant studies. In addition, methodological heterogeneity (e.g. in the reported effect sizes and the use of different covariates) in the individual studies complicated direct inter-study comparisons and hindered meta-analyses. This resulted in a relatively small number of studies (n=2) being included in the meta-analysis.

The individual studies included in this review also had inherent limitations. These studies included relatively small sample sizes, which may have limited statistical power to detect subtle but meaningful significant effects. The prevalence of mental disorders (e.g. externalising problem behaviour) in the included studies was also relatively low, which may have limited the power to detect significant associations (Suarez et al., 2018a). Thus, caution is needed when extrapolating these findings to larger study populations. Additionally, variations in participant recruitment methods, such as recruiting from schools (Cerveira de Baumont et al., 2021) compared to cohort studies (Tollenaar et al., 2021), may have introduced inconsistencies in diagnoses or phenotype classifications.

Finally, most of the studies in this review (n=3) made use of the Horvath epigenetic clock to measure EA deviation (Suarez et al., 2018a, Tollenaar et al., 2021, Cerveira de Baumont et al., 2021). This clock was previously reported to have reduced efficacy for estimating EA in children (Alisch et al., 2012, Higgins-Chen et al., 2020). Further, the Horvath clock (which was trained to predict chronological age) has been found to be less effective in detecting significant positive correlations between EAA and mental health outcomes (such as depression and schizophrenia), when compared to other epigenetic clocks trained for broader applications (such as predicting all-cause mortality) (Higgins-Chen et al., 2020). Thus, the performance of alternative epigenetic clocks (e.g. Pace of Aging methylation, PoA) in children and adolescents would warrant further exploration. Future studies could also benefit from comparing different epigenetic clocks for childhood-onset phenotypes to determine which clock is best suited for children.

In conclusion - notwithstanding these limitations - this systematic review highlights potential associations between EAA and adverse mental health outcomes in children and adolescents. While significant associations between EAA and internalising behaviour were described by individual studies, the meta-analysis did not confirm this finding. This underscores the need for larger and more well-powered studies, to enable further exploration of the utility of EA as early indicators of mental disorders in at-risk children.

## Data Availability

All data produced in the present work are contained in the manuscript

