## Appendix 1 for "Epigenetic age deviation and mental health in childhood and adolescence: A systematic review and meta-analysis"

**Table 1. Outline of search strategy tool**

| Spider tool |  | **Search terms** |
| --- | --- | --- |
| Sample | Children and adolescents | “young” OR”,” teen” OR “child*”, “youth”, “adolescent”, “young people” |
| Phenomenon of Interest | Epigenetic age (EA) deviation i.e., EA acceleration, EA deceleration, | “Epigenetic ageing”, “biological aging”, Epigenetic age acceleration”, |
| Design | All studies i.e., full-text articles |  |
| Evaluation | Development and mental health | Internalizing disorder, externalizing disorder, neurodevelopment disorder, developmental psychopathology  Cognitive development, behavioural disorders |
| Research type (qualitative, quantitative, mixed methods) | mixed methods, quantitative research |  |
